# Identifying Reasons for ACEI/ARB Non-Use in CKD Using Scalable Clinical NLP with Schema-Guided LLM Augmentation

**DOI:** 10.64898/2026.02.10.26346025

**Authors:** Mohammed Al-Garadi, Glenn Gobbel, L. Parker Gregg, Peter A. Richardson, Alex A. Herrera, Richard Noriega, Sheena Wydermyer, Jill Whitaker, Tina French, Michael McLemore, Dax M. Westerman, Sankar D. Navaneethan, Michael E. Matheny

## Abstract

**IMPORTANCE:** Although angiotensin-converting enzyme inhibitors (ACEIs) and angiotensin receptor blockers (ARBs) are recommended for people with chronic kidney disease (CKD), they remain underused. Barriers to adherence, such as adverse effects or patient refusal, are frequently embedded within unstructured clinical narratives and are therefore inaccessible to structured data analytics. Scalable natural language processing (NLP) approaches are needed to identify these barriers and support guideline-concordant care.

**OBJECTIVE:** To develop and evaluate an NLP model capable of identifying documented reasons for ACEI/ARB non-use within clinical notes of people with CKD in the Veterans Affairs (VA) healthcare system.

**DESIGN, SETTING, AND PARTICIPANTS:** This retrospective study analyzed electronic health record data from 2005 to 2024 including people aged 18 to 80 years with CKD, defined by an estimated glomerular filtration rate (eGFR) of 20–60 mL/min/1.73 m^2^ and presence of albuminuria, across multiple VA medical centers. NLP models were trained on 1,025 manually annotated notes and further augmented with 4,600 synthetic examples generated through schema-guided large language model prompting.

**MAIN OUTCOMES AND MEASURES:** The primary outcome was model performance in identifying notes containing at least one documented reason for ACEI/ARB non-use, evaluated using F1-score, precision, and recall. Secondary outcomes included model learning curve analyses and the effect of synthetic data augmentation on classification performance.

**RESULTS:** The most common documented reasons for ACEI/ARB non-use were acute kidney injury (29.6%), increased creatinine (12.4%), cough (11.2%), and hypotension-related symptoms (11.1%). Across modeling approaches, training with synthetic data augmentation improved detection of notes containing reasons for non-use. Performance gains were statistically significant across all models (McNemar test, P < .05), with the random forest model using Nomic embeddings achieving the highest performance (F1 score, 0.79; 95% CI, 0.68–0.90).

**CONCLUSIONS AND RELEVANCE:** We identified documented reasons for ACEI/ARB non-use (including both failures to initiate therapy and discontinuation after prior use) from unstructured text using an NLP method that does not require massive, expensive computing at inference time. By augmenting training data with schema-guided synthetic notes, we achieved robust, privacy-preserving performance within an NLP framework. This approach may support scalable clinical decision support systems to promote guideline-concordant prescribing.

## Introduction and Background

Chronic kidney disease (CKD) is a progressive condition characterized by a decline in kidney function over time. The renal protective effects of angiotensin-converting enzyme inhibitors (ACEIs) and angiotensin receptor blockers (ARBs) have led to their widespread use for slowing disease progression in people with CKD.^1^ Despite their established benefits, the prevalence of ACEI/ARB prescription in people with CKD varies across different populations and healthcare settings, and several studies have reported suboptimal utilization of these drugs.^2,3^ Improved utilization of ACEIs/ARBs in people with CKD has the potential to slow disease progression, reduce the risk of cardiovascular events, and improve patient outcomes.^4^

Discontinuation of these medicines is frequently due to adverse effects, such as acute kidney injury, angioedema, or hyperkalemia. Not reinitiating after intended temporary discontinuation contributes greatly to total underutilization of these agents.^5^ Identifying and assessing the factors contributing to underutilization in a given patient or population could help guide interventions to overcome these factors. Potential approaches at the healthcare system level include implementation of clinical decision support. However, a substantial challenge to implementing this is that the various factors that contribute to underutilization are often captured only within the unstructured text of patient clinical notes.^6^ The manual work required to identify and extract the relevant information with sufficient accuracy to reliably use the data for patient care is labor-intensive and requires significant clinical expertise.^7,8^

Natural language processing (NLP) has emerged as a promising tool to address these challenges by extracting relevant data from clinical notes.^9^ NLP techniques enable automated analysis of large volumes of unstructured data and might help to identify patient-specific factors,^10,11^ contraindications, and adverse events associated with ACEI/ARB non-use. NLP might aid in monitoring and detecting adverse events related to ACEIs/ARBs, providing valuable insights for optimizing therapy. It could also provide a comprehensive overview of a patient’s medical history and identify potential barriers to ACEI/ARB therapy.^5^ NLP algorithms can extract information regarding medication adherence, laboratory results, comorbidities, and concomitant medications, allowing for a more personalized approach to CKD management.^12^

Recent advances in large language models (LLMs), such as transformer-based architectures, have demonstrated strong general language understanding and the ability to perform zero-shot (no task-specific examples provided), one-shot (a single example provided), and few-shot (a small number of examples provided) classification across a range of biomedical tasks.^13,14^ However, deploying LLMs in clinical settings remains challenging because of inference latency, computational cost, and constraints on protected health information.^15,16^ To balance these practical constraints with the strengths of LLMs, we developed a hybrid framework.

With a goal of addressing the needs and challenges noted above, we sought to develop and evaluate an NLP-based tool capable of automatically and rapidly analyzing clinical notes from Veterans with CKD and identifying reasons for not prescribing or for discontinuing ACEIs and ARBs. This included deployment of a hybrid system that integrated schema-guided manual annotation with synthetic clinical notes generated by a structured LLM prompting approach to augment a manually annotated corpus and train traditional NLP classifiers. This strategy enables robust learning of diverse clinical documentation patterns, particularly for rare or underrepresented barriers to ACEI/ARB use, without relying on costly or difficult-to-deploy generative models. We hypothesized that this approach would outperform traditional NLP and offer a more reliable, scalable deployment than LLMs, which are limited by high costs, non-deterministic outputs, and PHI constraints.^16-19^

## Methods

### Analytical Workflow Overview

To study ACEI/ARB non-use in CKD, we developed a structured NLP pipeline that integrates expert-driven annotation with schema-guided LLM-based data augmentation. Manual review was used to define and quantify clinically documented reasons for non-use, and NLP models were trained to identify clinical notes containing such documentation at scale. This approach enables efficient identification of patients with documented or undocumented gaps in guideline-recommended therapy and supports scalable downstream clinical decision support.

The pipeline is summarized in Figure 1. Briefly, we first defined standardized annotation guidelines and manually labeled a reference corpus of CKD-related clinical notes. Schema-guided prompt engineering was then applied to a large language model to generate additional positive and negative examples. Expert- and LLM-generated notes were split into training (70%), validation (10%), and test (20%) sets. Models were trained on either the original or augmented data using classic word-level representations with logistic regression, random forest, or XGBoost, as well as fine-tuned transformer models. Final evaluation on the held-out test set employed precision, recall, F1-score with 95% CI, McNemar’s test, and learning-curve analysis.

**Figure 1.**
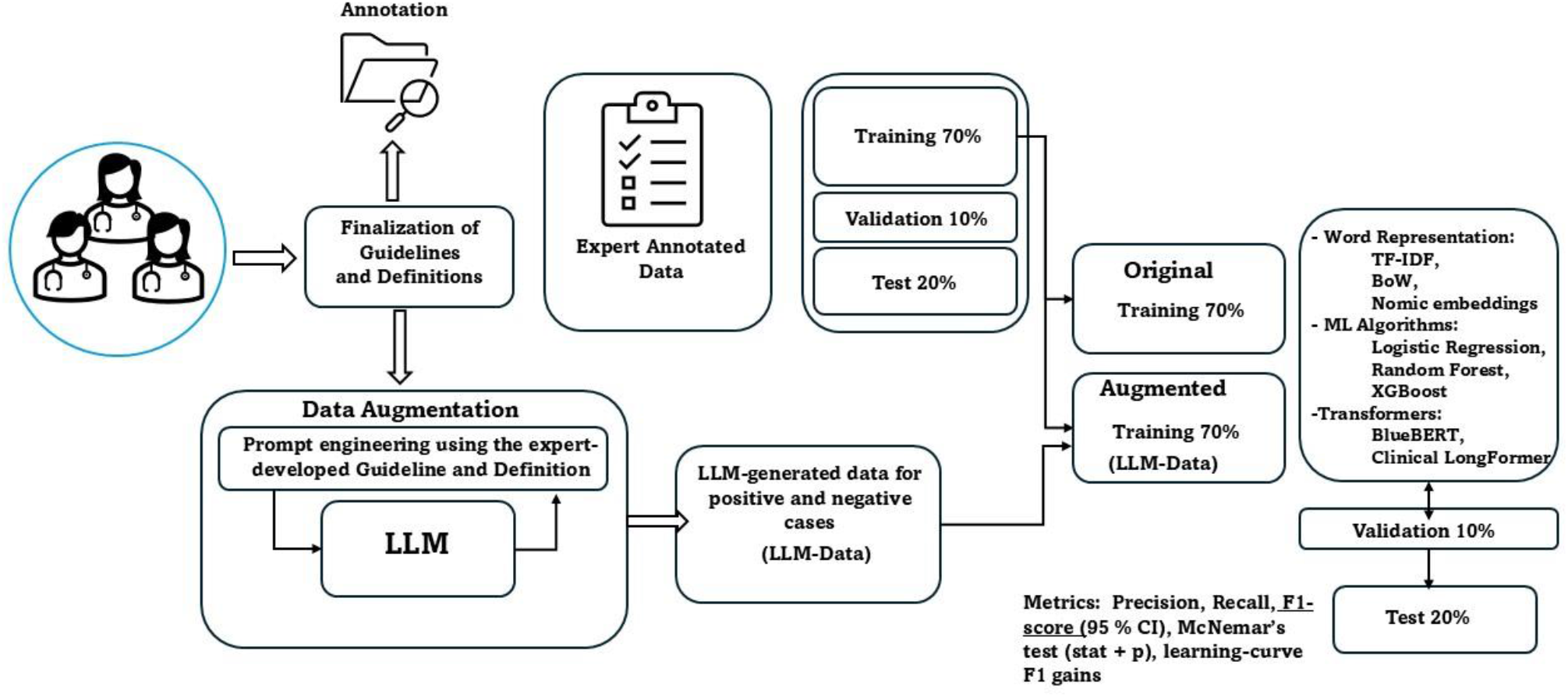
Workflow for Developing and Evaluating NLP Models to Detect Reasons for ACEI/ARB Non-Use in CKD.

### Study Design and patient cohort

We conducted a retrospective analysis of clinical notes from VA patients with CKD. The cohort included patients aged 18–80 years with albuminuria and an estimated glomerular filtration rate (eGFR) of 20–60 mL/min/1.73 m^2^, measured on two occasions at least 90 days apart between January 1, 2005, and December 31, 2024. Patients were identified using a CKD database developed by our team based on VA structured datasets, which has also been applied to other health systems using electronic medical records. Among these patients, those prescribed ACEI or ARB therapy were identified through VA pharmacy records. For analyses focused on initiation following CKD diagnosis, we restricted the cohort to patients with outpatient ACEI/ARB prescriptions after the CKD index date and no prior record of ACEI or ARB dispensation within the VA system, thereby defining an incident-user population.

### Data Collection

Data for this study was obtained from the VA healthcare system’s electronic health record (EHR) database using resources provided by VA Office of Research and Development VA Informatics and Computing Infrastructure (VINCI) resource center. Specifically, we used: (a) inpatient and outpatient SQL datasets to capture patient-level demographics and comorbidities, based on Current Procedural Terminology (CPT) codes and International Classification of Disease, Clinical Modification, Ninth and Tenth revisions (ICD-9-CM and ICD-10-CM, respectively) diagnosis and procedure codes (ICD-9-CM through September 30, 2015; ICD-10-CM on or after October 1, 2015); (b) the VA Managerial Cost Accounting System Laboratory Results database for serum creatinine and other relevant laboratory data; (c) the Corporate Data Warehouse Production Outpatient Pharmacy domain for detailed prescription information; and (d) the VA Vital Status and Beneficiary Identification Records Locator Subsystem (BIRLS) files for mortality data. This dataset provided a representative sample of people with CKD across diverse populations and healthcare settings.

### Note Selection

Clinical notes used for NLP tool development and evaluation were derived from the EHRs of patients within the cohort described above. To enrich the corpus for notes likely to contain reasons for ACEI/ARB non-use, we restricted selection to documents anchored to key clinical dates and those linked to allergy/adverse event tables. Notes were further limited to those authored by pharmacists, nurse practitioners, physicians, and physician assistants, and to documents coded within the VA system with subject matter domains or titles relevant to ACEI/ARB non-use. Clinical providers (SK, MM, SV) and nurse annotators (JW, TF) from the project team manually reviewed all subject domains and titles to confirm relevance.

### Annotation Schema

We generated an annotation schema to define concepts of interest within the clinical, unstructured text notes that related to potential reasons for ACEI/ARB non-use. The schema definitions provided guidelines for reviewers regarding the phrases that should be annotated and would be used for NLP tool development and evaluation. The schema was developed using an iterative process that started with clinical experts on the team (SN, MM, SV) generating a list of specific concepts that are commonly associated with non-use, such as dry cough, hyperkalemia, and angioedema. Two clinical informatics nurses (JW and TF) then reviewed a preliminary subset of approximately 25 notes and annotated any concepts within the aforementioned list and also noted any potentially relevant concepts not included in the list. Annotations consisted of marking sequences of one or more tokens within the text corresponding to a concept of interest, where a token is defined as a unit of meaning in the text, which is often a word but may also be other character sequences such as numbers, dates, and units of measure. The specific concept list was updated by the clinical experts and annotators and the process repeated until no further schema refinements were required.

The schema contained five broad concepts of interest, including 1) explicit mentions of ACEIs and ARBs, 2) well-recognized adverse effects of these agents, including cough, angioedema, increased creatinine, hyperkalemia, and hypotension, 3) non-specific allergies and general intolerance 4) patient refusal, and 5) a perceived lack of efficacy of these agents by provider.^20,21^ Annotators also assigned a positive or negative assertion value to all annotated concepts. After completing the annotation of all concepts of interest, reviewers classified the note according to whether it contained one or more reasons for a provider not prescribing an ACEI or ARB. Inter-rater agreement between the two primary annotators was assessed using Cohen’s kappa statistic.^22^

### Model Development

We developed and systematically evaluated machine learning models for clinical note classification, optimized for deployment in resource-constrained environments. Model selection prioritized effectiveness in clinical use, computational efficiency, and scalability for integration into real-world healthcare systems.^23-25^ This study was conducted under the hypothesis that traditional NLP approaches can perform competitively across diverse clinical text classification tasks when provided with sufficiently large and heterogeneous training data.

#### Text Representation Techniques

Clinical notes were transformed into numerical features using three methods: (1) Term Frequency-Inverse Document Frequency (TF-IDF),^26^ assigning weights to words based on their relative importance in each document compared to the entire dataset; (2) Bag-of-Words (BoW), quantifying word frequencies without capturing context;^27^ and (3) Transformer-based sentence Nomic embeddings.^28^

#### Machine Learning Algorithms

The study evaluated five machine learning algorithms: (1) Logistic Regression,^29^ (2) Random Forest,^30^ and (3) XGBoost,^31^ each combined individually with three distinct text representations: TF-IDF, BoW, and Nomic embeddings.^28^ Two transformer-based deep learning models were also evaluated: (4) Clinical Longformer,^32^ pretrained specifically on clinical notes to capture nuanced clinical language, and (5) BlueBERT,^33,34^ pretrained on biomedical texts to capture nuanced biomedical language.

#### Training and Evaluation

The dataset was partitioned into training (70%), validation (10%), and test (20%) sets. Models were evaluated on the held-out test set using class-specific sensitivity, specificity, positive predictive value, negative predictive value, and F1-score, with 95% confidence intervals, across all text representations and algorithm combinations.

#### Statistical Evaluation and Performance Comparison

Model performance was presented alongside 95% confidence intervals obtained through bootstrap resampling. McNemar’s test compared performances between models trained on original and augmented data, with statistical significance defined at p < 0.05.

### Data Augmentation via Large Language Models

#### Synthetic Note Generation Using Annotation Schema

To supplement the limited number of annotated clinical notes and to ensure adequate representation of rare or ambiguous documentation scenarios, we used a structured annotation schema to guide the generation of synthetic notes via GPT-4.0.^35^ The schema was developed iteratively by clinical experts and clinical informatics nurse annotators and operationalized core documentation patterns related to ACEI/ARB non-use, including medication mentions, adverse effects, intolerance or allergy, patient refusal, and provider-documented perceptions of inefficacy. Synthetic note generation preserved span-level concept structure and assertion status (positive or negative) consistent with the manual annotation process (see Annotation Schema). The annotation schema was derived exclusively from manual review of real clinical notes by clinical nurse annotators.

To mitigate hallucination risk, schema-guided prompting was used to constrain synthetic note generation to predefined, clinically validated documentation patterns. Synthetic notes were generated to match real clinical notes in length, narrative structure, and volume of clinical information. Each synthetic note contained no more than one documented reason for ACEI/ARB non-use and included additional clinical content such as symptoms, laboratory results, comorbid conditions, and concurrent medications. Prompt development was iterative and grounded in the annotation schema and review of model classification errors, with prompts explicitly specifying allowable concept categories, assertion polarity, and inclusion of documentation patterns known to challenge automated classification

The synthetic notes were incorporated into the training set to increase concept coverage and improve model robustness, particularly for underrepresented or ambiguous cases.

#### Error Typology and Model Failure Analysis

To improve performance on underrepresented cases, we conducted detailed error analysis using validation predictions from the baseline model. This analysis identified concept categories frequently missed or misclassified, such as angioedema, patient refusal, and perceived ineffectiveness. Targeted synthetic notes were then generated to supplement these rare cases. GPT-4.0 prompts were refined to produce semantically diverse, schema-aligned examples representing these specific scenarios.

All decisions related to synthetic note generation, volume, content focus, and prompt structure, were informed exclusively by training and validation data. The independent test set was withheld throughout this process and used only for final model evaluation to ensure unbiased performance assessment.

Error analysis and subsequent schema-guided synthetic note generation were informed by training and validation data only and were used to improve the representation of underdocumented clinical documentation patterns. Model performance was subsequently evaluated on a held-out test set that was not used for model training or synthetic data generation, enabling assessment of generalization to unseen clinical notes despite differences in data composition.

#### Determining Counts of Synthetic Note Generation via Learning Curve

To optimize the number of synthetic clinical notes incorporated into model training, we implemented a learning curve–based approach using the validation set. Models were initially trained on human-annotated notes alone and evaluated on the validation set to establish a baseline. Synthetic notes were then added incrementally in predefined batches. After each addition, model performance was re-evaluated on the validation set using F1-score, recall, and concept-level precision. The point at which additional synthetic data failed to yield meaningful performance gains was used to determine the final augmentation volume.

## Results

### Annotation

From 1,025 annotated documents, distinct reasons for discontinuation of ACEI or ARB therapy were identified. Acute kidney injury was the most frequently cited reason (29.6%), followed by increased creatinine (12.4%), cough (11.2%), and hypotension-related symptoms (11.1%) (Figure 2). Additional reasons included non-specific allergy, hyperkalemia, and a small number of behavioral factors, such as patient non-adherence. Rare entries reflected overlapping adverse events, including combinations such as acute kidney injury with hyperkalemia or hypotension. These annotations illustrate the diverse clinical contexts influencing decisions to discontinue treatment. Inter-annotator agreement for the classification of reasons for non-use was substantial, yielding a Cohen’s kappa of >0.80.

**Figure 2.**
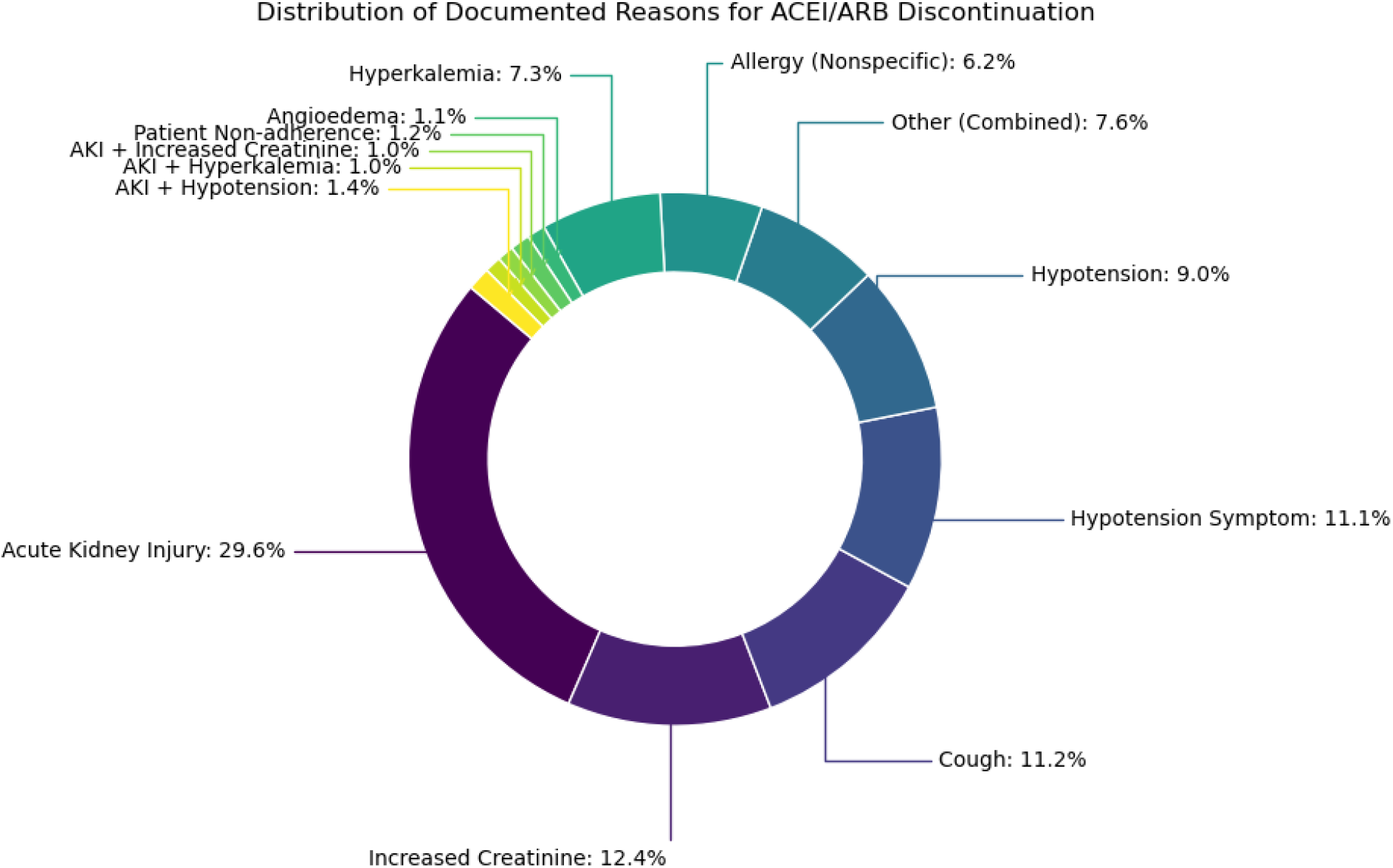
Distribution of Documented Reasons for ACEI or ARB Discontinuation Among 1,025 Annotated Cases.

### Synthetic Data Generation for Category Augmentation

A total of 4,600 synthetic clinical note–style examples were generated across 23 predefined categories representing reasons for ACEI/ARB discontinuation, intolerance, or mention. Each category was augmented with 200 examples, using a prompt-based approach informed by definitions, sample phrases, and context keywords (Table 1). These categories included both clinical events (e.g., *Acute Kidney Injury, Angioedema, Cough*) and behavioral or contextual factors (e.g., *Patient Refusal, Non-Adherence, Other Causes of Non-Use*). This corpus supports downstream model training for improved identification and classification of ACEI/ARB-related documentation in clinical text.

**Table 1.**
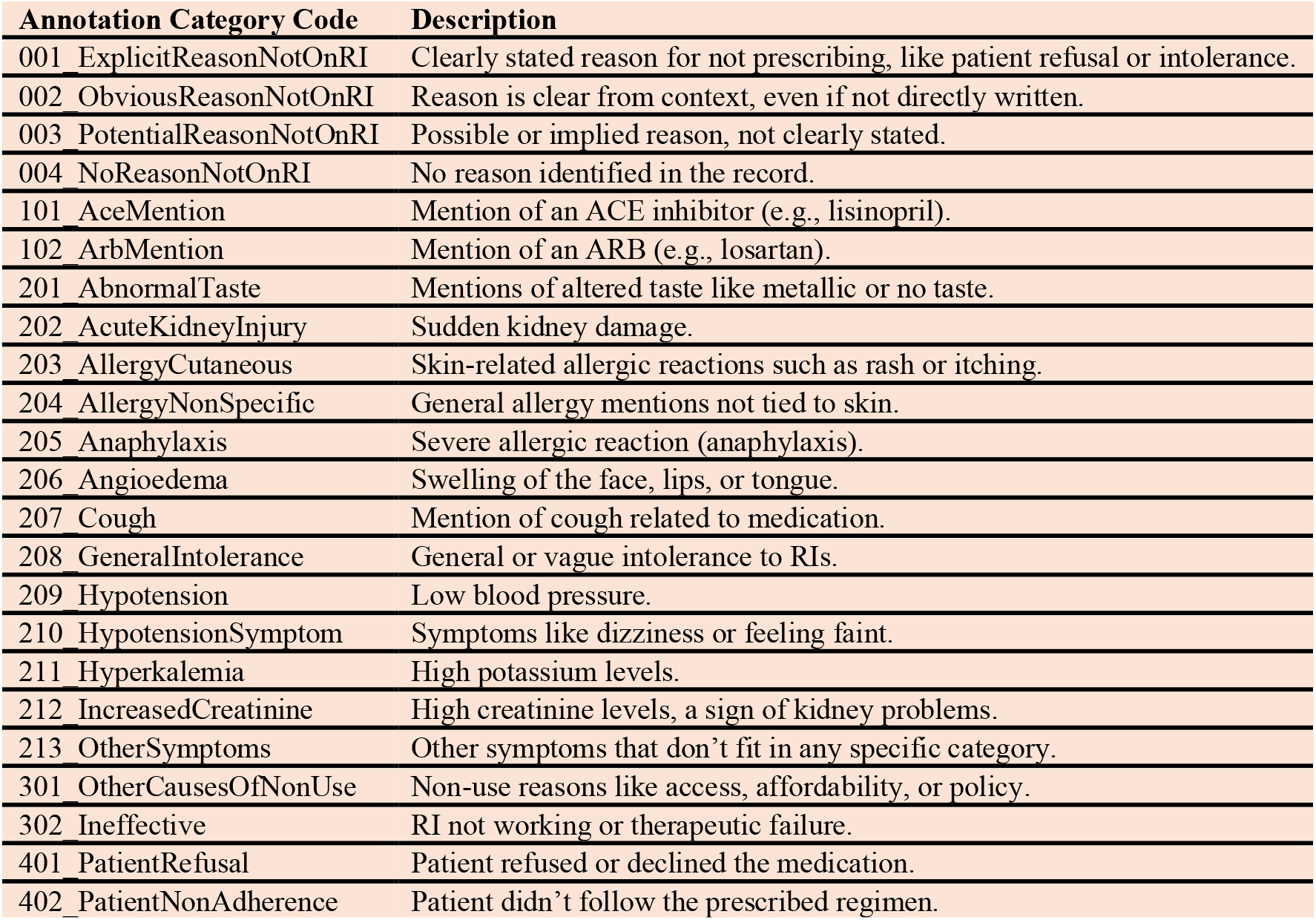
Schema Categories and Descriptions for ACEI/ARB Non-Use and Related Mentions in Clinical Notes.

### Models’ Performance Across Representations

Given substantial class imbalance, performance interpretation focused on the minority ‘Reason’ class, with observed improvements driven primarily by increased precision for clinically relevant reason detection rather than changes in majority-class (‘No_reason’) performance.

The evaluation of model performance across TFIDF, Bag-of-Words, Nomic, and Transformer representations, with and without augmented data, revealed distinct outcomes for “No_reason” and “Reason” classes. Metrics are reported as precision, recall, and F1-scores with 95% confidence intervals (CI), reflecting high statistical rigor.

With TF-IDF representation (Table 2), data augmentation increased Logistic Regression performance for the “Reason” class, yielding an F1-score of 0.66 (95% CI, 0.53–0.79), representing an approximately 8% improvement compared with training on original data alone. Augmentation also improved XGBoost performance, increasing the F1-score to 0.73 (95% CI, 0.66–0.80), corresponding to a ∼7% increase. Random Forest models demonstrated the largest augmentation-associated gain, with F1-score increasing from 0.50 (95% CI, 0.43–0.57) without augmentation to substantially higher performance with augmented training data, reflecting an approximate 36% improvement.

**Table 2.** Performance of Models Using TFIDF Representation with and without Augmented Data.

| Classifier | Data | Sensitivity<br>(Recall-Reason) | Specificity<br>(Recall-No_reason) | PPV<br>(Precision-Reason) | NPV<br>(Precision-No_reason) | F1-Reason | F1-No_reason |
| --- | --- | --- | --- | --- | --- | --- | --- |
| **Logistic Regression** | *Original* | 0.84<br>(0.74–0.94) | 0.96<br>(0.94–0.98) | 0.49<br>(0.39–0.59) | 0.99<br>(0.98–1.00) | 0.61<br>(0.48–0.74) | 0.98<br>(0.96–0.99) |
|  | *Augmented* | 0.63<br>(0.55–0.71) | 0.99<br>(0.98–1.00) | 0.71<br>(0.59–0.83) | 0.99<br>(0.98–1.00) | 0.66<br>(0.53–0.79) | 0.99<br>(0.98–1.00) |
| **XGBoost** | *Original* | 0.79<br>(0.67–0.91) | 0.98<br>(0.96–0.99) | 0.60<br>(0.47–0.73) | 0.99<br>(0.98–1.00) | 0.68 (0.61–0.75) | 0.99<br>(0.98–1.00) |
|  | *Augmented* | 0.74<br>(0.63–0.85) | 0.99<br>(0.98–1.00) | 0.73<br>(0.61–0.85) | 0.99<br>(0.98–1.00) | 0.73<br>(0.66–0.80) | 0.99<br>(0.98–1.00) |
| **Random Forest** | *Original* | 0.84<br>(0.74–0.94) | 0.94<br>(0.92–0.96) | 0.36<br>(0.28–0.44) | 0.99<br>(0.98–1.00) | 0.50<br>(0.43–0.57) | 0.97<br>(0.96–0.98) |
|  | *Augmented* | 0.63<br>(0.50–0.76) | 0.99<br>(0.98–1.00) | 0.74<br>(0.62–0.86) | 0.99<br>(0.97–1.00) | 0.68<br>(0.55–0.81) | 0.99<br>(0.98–1.00) |

For Bag-of-Words representation (Table 3), data augmentation substantially improved the detection of the “Reason” class across all classifiers. Logistic Regression showed a marked performance gain, with the F1-score increasing to 0.81 (95% CI, 0.70–0.92), representing an approximately 29% improvement over training on original data alone. XGBoost performance also improved with augmentation, reaching an F1-score of 0.72 (95% CI, 0.61–0.84), corresponding to a ∼16% increase. Random Forest models demonstrated the largest augmentation-associated benefit, with F1-score increasing from 0.52 (95% CI, 0.39–0.65) without augmentation to 0.73 (95% CI, 0.60–0.86) with augmented data, reflecting an approximate 40% improvement. Performance for the No_reason class remained consistently high across models.

**Table 3.** Performance of Models Using Bag-of-Words Representation with and Without Augmented Data.

| Classifier | Data | Sensitivity<br>(Recall-Reason) | Specificity<br>(Recall-No_reason) | PPV<br>(Precision-Reason) | NPV<br>(Precision-No_reason) | F1-Reason | F1-No_reason |
| --- | --- | --- | --- | --- | --- | --- | --- |
| **Logistic Regression** | *Original* | 0.79<br>(0.71–0.87) | 0.97<br>(0.96–0.99) | 0.54<br>(0.43–0.65) | 0.99<br>(0.98–1.00) | 0.63<br>(0.56–0.70) | 0.98<br>(0.97–0.99) |
|  | *Augmented* | 0.79<br>(0.71–0.87) | 0.99<br>(0.99–1.00) | 0.83(0.75–0.91) | 0.99<br>(0.98–1.00) | 0.81<br>(0.70–0.92) | 0.99<br>(0.99–1.00) |
| **XGBoost** | *Original* | 0.74<br>(0.66–0.82) | 0.98<br>(0.96–0.99) | 0.53<br>(0.41–0.65) | 0.99(0.98–1.00) | 0.62<br>(0.53–0.71) | 0.98<br>(0.97–0.99) |
|  | *Augmented* | 0.72<br>(0.61–0.85) | 0.99<br>(0.98–1.00) | 0.73<br>(0.66–0.80) | 0.99<br>(0.98–1.00) | 0.72<br>(0.61–0.84) | 0.99<br>(0.98–1.00) |
| **Random Forest** | *Original* | 0.89<br>(0.81–0.97) | 0.94<br>(0.92–0.96) | 0.37<br>(0.27–0.47) | 0.99<br>(0.98–1.00) | 0.52(<br>0.39–0.65) | 0.97<br>(0.96–0.98) |
|  | *Augmented* | 0.73<br>(0.63–0.83) | 0.99<br>(0.98–1.00) | 0.74<br>(0.64–0.84) | 0.99<br>(0.98–1.00) | 0.73<br>(0.60–0.86) | 0.99<br>(0.98–1.00) |

With Nomic representation (Table 4), data augmentation led to consistent relative improvements in the identification of the “Reason” class across all classifiers. Logistic Regression performance increased to an F1-score of 0.73 (95% CI, 0.60–0.86), representing an approximately 22% improvement over training on original data alone. XGBoost showed a more modest gain with augmentation, reaching an F1-score of 0.71 (95% CI, 0.59–0.83), corresponding to a ∼9% increase. Random Forest models demonstrated the strongest augmentation-associated benefit under this representation, with F1-score improving from 0.63 (95% CI, 0.53–0.73) without augmentation to 0.79 (95% CI, 0.68–0.90) with augmented data, reflecting an approximate 25% improvement.

**Table 4.** Performance of Models Using Nomic Representation with and without Augmented Data.

| Classifier | Data | Sensitivity<br>(Recall-Reason) | Specificity<br>(Recall-No reason) | PPV<br>(Precision-Reason) | NPV<br>(Precision-No reason) | F1-Reason | F1-No_reason |
| --- | --- | --- | --- | --- | --- | --- | --- |
| **Logistic Regression** | *Original* | 0.79<br>(0.67–0.91) | 0.97<br>(0.95–0.98) | 0.48<br>(0.37–0.59) | 0.99<br>(0.98–1.00) | 0.60<br>(0.52–0.68) | 0.98<br>(0.97–0.99) |
|  | *Augmented* | 0.79<br>(0.71–0.87) | 0.99<br>(0.97–1.00) | 0.68<br>(0.56–0.80) | 0.99<br>(0.98–1.00) | 0.73<br>(0.60–0.86) | 0.99<br>(0.98–0.99) |
| **XGBoost** | *Original* | 0.79<br>(0.69–0.89) | 0.98<br>(0.96–0.99) | 0.56<br>(0.47–0.65) | 0.99<br>(0.98–1.00) | 0.65<br>(0.54–0.76) | 0.98<br>(0.97–0.99) |
|  | *Augmented* | 0.79<br>(0.69–0.89) | 0.98<br>(0.97–0.99) | 0.65<br>(0.57–0.73) | 0.99<br>(0.98–1.00) | 0.71<br>(0.59–0.83) | 0.99<br>(0.98–0.99) |
| **Random Forest** | *Original* | 0.84<br>(0.75–0.93) | 0.97<br>(0.95–0.98) | 0.50<br>(0.43–0.57) | 0.99<br>(0.98–1.00) | 0.63<br>(0.53–0.73) | 0.98<br>(0.97–0.99) |
|  | *Augmented* | 0.68<br>(0.60–0.76) | 1.00<br>(0.99–1.00) | 0.93<br>(0.84–0.96) | 0.99<br>(0.98–1.00) | 0.79<br>(0.68–0.90) | 0.99<br>(0.99–1.00) |

For Clinical Longformer (Table 5), augmentation increased the F1-score for the “Reason” class to 0.73 (95% CI, 0.70–0.78), corresponding to an approximately 9% relative improvement compared with training on original data alone. BlueBERT showed a larger augmentation-associated gain, with F1-score increasing to 0.78 (95% CI, 0.72–0.90), reflecting an approximate 15% improvement.

**Table 5.** Performance of Finetuned Transformer Models with and Without Augmented Data.

| Classifier | Data | Sensitivity<br>(Recall-Reason) | Specificity<br>(Recall-No reason) | PPV<br>(Precision-Reason) | NPV<br>(Precision-No reason) | F1-Reason | F1-No_reason |
| --- | --- | --- | --- | --- | --- | --- | --- |
| Clinical Longformer | *Original* | 0.74<br>(0.70–0.78) | 0.98<br>(0.96–0.99) | 0.60<br>(0.55–0.65) | 0.99<br>(0.98–1.00) | 0.67<br>(0.61–0.74) | 0.98<br>(0.97–0.99) |
|  | *Augmented* | 0.80<br>(0.71–0.87) | 0.98<br>(0.97–0.99) | 0.67<br>(0.59–0.73) | 0.99<br>(0.98–1.00) | 0.73<br>(0.70–0.78) | 0.99<br>(0.98–0.99) |
| BlueBERT | *Original* | 0.68<br>(0.64–0.72) | 0.97<br>(0.95–0.98) | 0.65<br>(0.63–0.70) | 0.99<br>(0.98–1.00) | 0.68<br>(0.65–0.72) | 0.98<br>(0.97–0.99) |
|  | *Augmented* | 0.68<br>(0.60–0.76) | 1.00<br>(0.99–1.00) | 0.92<br>(0.83–0.95) | 0.99<br>(0.99–1.00) | 0.78<br>(0.72–0.90) | 0.99<br>(0.99–1.00) |

#### Statistical Comparison of Top Models for the ‘Reason’ Class

The top three models for the “Reason” class using augmented data (Table 6) were Random Forest (Nomic, F1-score: 0.79), BlueBERT (Transformer, F1-score: 0.78), and Clinical Longformer (Transformer, F1-score: 0.73). McNemar’s tests demonstrated statistically significant differences in paired predictions between models trained with and without data augmentation. The random forest model (χ^2^ = 3.0; P = .008), BlueBERT (χ^2^ = 2.76; P = .011), and Clinical Longformer (χ^2^ = 2.15; P = .032) each showed significant improvement with augmentation (P < .05). McNemar’s tests confirmed statistically significant differences in predictions between models trained on original versus augmented data. Random Forest showed a test statistic of 3.000 (p = 0.008), BlueBERT 2.758 (p = 0.011), and Clinical Longformer 2.145 (p = 0.032), all indicating significance at p < 0.05. These findings suggest that data augmentation led to meaningful improvements in model performance for all top models when detecting reasoning-related information.

**Table 6.** Performance of Top 3 Models on the “Reason” Class Using Augmented Data.

| Model | Representation | Precision (95% CI) | Recall (95% CI) | F1-Score (95% CI) |
| --- | --- | --- | --- | --- |
| **Random Forest** | **Nomic** | **0.93 (0.84–0.96)** | **0.68 (0.60–0.76)** | **0.79 (0.68–0.90)** |
| BlueBERT | Transformer | 0.92 (0.83–0.95) | 0.68 (0.60–0.76) | 0.78 (0.72–0.90) |
| Clinical Longformer | Transformer | 0.67 (0.59–0.73) | 0.80 (0.71–0.87) | 0.73 (0.70–0.78) |

#### Learning Curve Analysis

Model performance improved with increasing volumes of stratified synthetic notes. Random Forest-Nomic demonstrated the largest absolute gain, with F1-score increasing from 0.63 without augmentation to 0.79 at 100% augmentation. BlueBERT improved from 0.68 to 0.775, with most gains observed by 50% augmentation. Clinical Longformer showed a more gradual increase, reaching its peak F1-score of 0.73 at 75% augmentation. Performance for both BlueBERT and Clinical Longformer plateaued beyond 75%, whereas Random Forest-Nomic continued to improve through full augmentation (See Figure 3).

**Figure 3.**
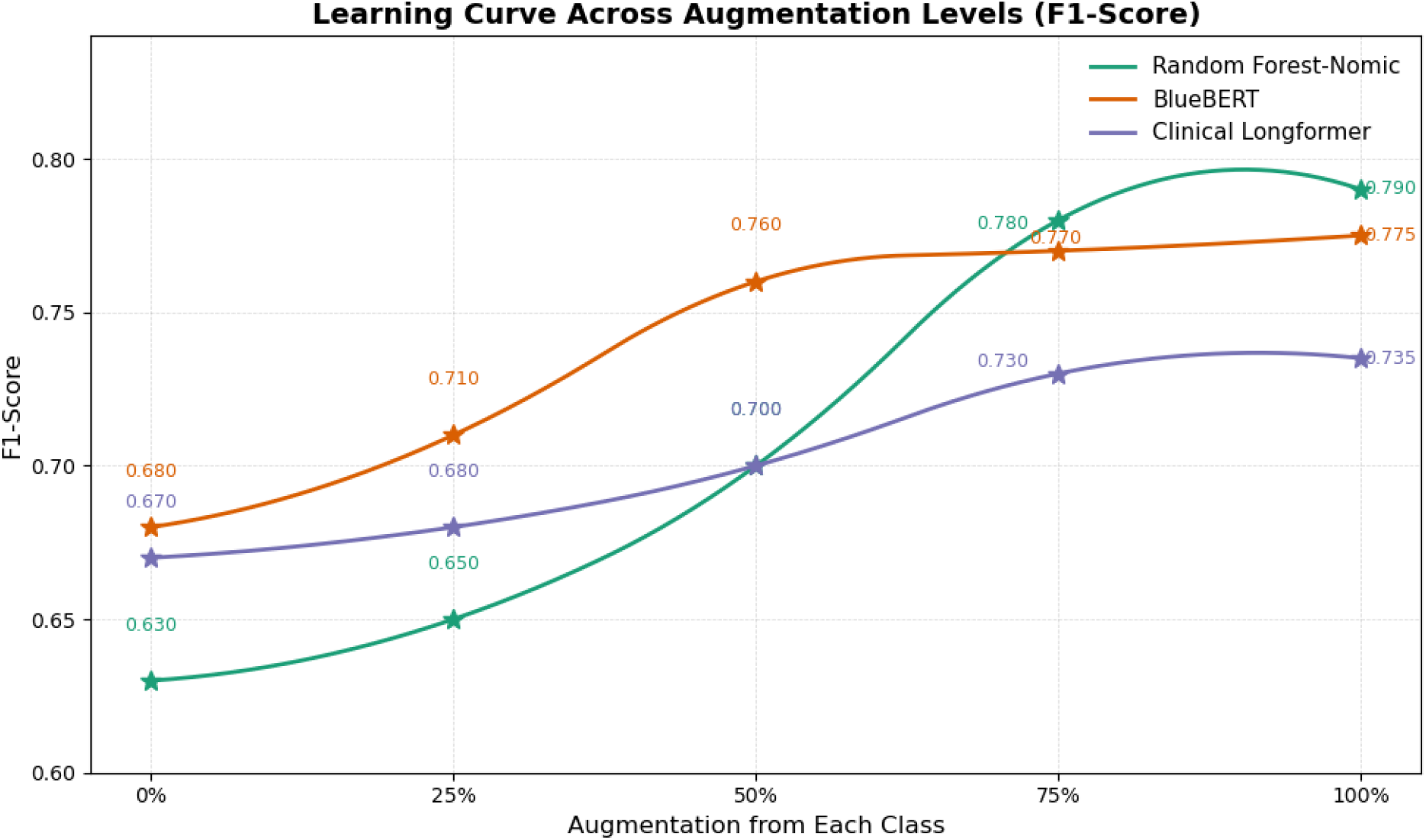
Impact of Data Augmentation on Model Performance: A Comparative Analysis of F1-Scores Across Augmentation Levels.

## Discussion

In this retrospective analysis of VA clinical data, we developed and evaluated a scalable hybrid NLP framework capable of extracting clinically meaningful information from unstructured notes to identify reasons for ACEI/ARB non-use among patients with CKD. Our findings address a critical and persistent gap in CKD management—suboptimal utilization of guideline-recommended therapies due to clinical, behavioral, or documentation-related barriers.^5,36-39^ This hybrid framework establishes a significant contribution by utilizing LLMs as a high-fidelity data engine to empower lightweight, privacy-preserving models that are feasible to deploy within existing clinical firewalls.

Our results confirm that reasons for non-use of ACEIs/ARBs are frequently embedded within narrative clinical documentation, making them inaccessible to conventional structured-data analysis. NLP, when trained using clinically grounded annotation schemas and augmented with high-quality synthetic examples, demonstrated strong performance in detecting diverse scenarios ranging from acute adverse reactions (e.g., angioedema, hyperkalemia) to patient refusal and provider perception of inefficacy. This aligns with earlier findings on the variability and complexity of treatment discontinuation in CKD^40^ but uniquely extends the literature by operationalizing these insights through automated, scalable methods.

A key innovation in this study was the use of LLM-generated synthetic notes, guided by a clinically validated schema, to augment rare or ambiguous classes. This approach improved model performance across all machine learning paradigms tested, with particularly strong gains observed in transformer-based models and random forest classifiers using Nomic embeddings. Beyond performance, our synthetic data generation framework also enhanced model robustness by incorporating semantically challenging edge cases, often overlooked in human-annotated corpora.

Compared to traditional NLP approaches that rely on rule-based heuristics or static ontologies, our method demonstrates superior adaptability and coverage of diverse clinical language. Rule-based systems often struggle to capture the nuanced,^41,42^ evolving ways in which clinicians document reasons for medication non-use, particularly when phrased indirectly or embedded within complex narratives.^7,43,44^ By contrast, our pipeline combines schema-guided annotation with synthetic data augmentation, enabling learning from edge cases and ambiguous contexts that static systems routinely miss. Moreover, while LLMs such as GPT-4 offer strong performance in few-shot settings, they present practical limitations in healthcare, including high inference costs, opacity in decision-making, and institutional constraints on protected health information handling.^45^ Our hybrid framework offers a scalable alternative: utilizing the generative reasoning of LLMs to create ‘training curricula’ for smaller, secure, and cost-effective models.^46^ This balance between generalization and operational feasibility positions our approach as a practical advancement in clinical NLP.

The learning curve analyses further support the efficiency of this approach, highlighting performance gains that plateau after strategic volumes of augmentation, insights that can guide future NLP development efforts in low-resource clinical domains.

### Limitations

This study has several important limitations. First, while we utilized GPT-4 for synthetic data generation, we did not employ it for direct inference. This was a deliberate design choice to prioritize the development of privacy-preserving, local models, though future work may benchmark these against secure, on-premise LLM deployments. As a result, we could not perform head-to-head benchmarking against state-of-the-art LLMs that may demonstrate higher performance in zero- or few-shot scenarios. Second, our dataset was drawn exclusively from the VA healthcare system, which may limit generalizability to other settings, particularly those with different patient populations or documentation styles. Third, while synthetic note generation significantly improved model robustness, errors persisted in distinguishing vague or overlapping concepts (e.g., allergy vs. intolerance). Lastly, the retrospective design introduces potential bias due to missing or inconsistently documented information. Future prospective validation in diverse clinical environments is warranted.

### Future work

Future work should explore the integration of this NLP tool into clinical care pathways, enabling real-time identification of patients at risk for ACEI/ARB underuse. Prospective validation across health systems with diverse populations will also be critical for confirming generalizability. Moreover, expanding the annotation schema to include social and structural factors documented in free text, such as cost concerns or access barriers, may uncover additional targets for intervention.

## Conclusion

This study demonstrates that tailored NLP systems can accurately identify reasons for ACEI/ARB non-use in patients with CKD, addressing a key challenge in leveraging unstructured clinical data for real-world decision support. Through schema-guided annotation and synthetic note augmentation, we achieved high model performance without relying on large-scale generative models or extensive computing infrastructure. While we were unable to directly evaluate or deploy high-parameter LLMs such as GPT-4 due to protected health information and resource constraints, our findings support the feasibility of using structured, domain-informed approaches to build clinically useful NLP tools in real-world environments. Future work should explore secure, scalable integration of more advanced LLMs in clinical settings and extend validation to diverse healthcare systems.

## Data Availability

The data that support the findings of this study were obtained from the United States Department of Veterans Affairs and are not publicly available due to federal data security and privacy requirements. Access to these data may be granted to qualified investigators with appropriate approvals through the VA research data environment. The annotation schema, and analysis code are available from the corresponding author upon reasonable request and subject to institutional approvals.

## Acknowledgments

This work was supported by the U.S. Department of Veterans Affairs, Veterans Health Administration, Office of Research and Development. The authors acknowledge the VA-VINCI for providing access to data and computational resources used in this study — VINCI: Putting Data to Work for Veterans. The contents do not represent the views of the U.S. Department of Veterans Affairs or the United States Government.

